# Incidence, prevalence, and survival of colorectal cancer in the United Kingdom from 2000-2021: a population-based cohort study

**DOI:** 10.1101/2024.07.12.24310284

**Authors:** Patricia Pedregal-Pascual, Carlos Guarner-Argente, Eng Hooi Tan, Asieh Golozar, Talita Duarte-Salles, Andreas Weinberger Rosen, Antonella Delmestri, Wai Yi Man, Edward Burn, Daniel Prieto Alhambra, Danielle Newby

**Author notes:** **Corresponding author(s):** Professor Daniel Prieto Alhambra, Centre for Statistics in Medicine, Nuffield Department of Orthopaedics, Rheumatology and Musculoskeletal Sciences, University of Oxford, Oxford, UK. These authors contributed equally to this work (Patricia Pedregal-Pascual, Carlos Guarner-Argente).

## Abstract

**Background:** The management of colorectal cancer (CRC) is evolving, with advances in screening and treatment. We leveraged population-based data to generate up-to-date UK estimates of age and sex-specific incidence and prevalence and overall survival for the period 2000-2021.

**Methods:** We analysed nationally representative primary care records from Clinical Practice Research Datalink (CPRD) GOLD, and replicated in CPRD Aurum. We calculated incidence rates, prevalence, and short- and long-term survival stratified by age, sex, and diagnosis year.

**Results:** Unadjusted prevalence increased in the study period, from 15.6 to 46.4/10,000. Overall incidence was 61.5/100,000 person years, increasing in 2000-2011 to drop slightly in 2011-2014, and then plateauing. In contrast, early-onset CRC raised uninterruptedly throughout the study period, from 8.33 to 19.07/100,000 person-years.

Overall survival was 78.3%, 51.4% and 38.5% at 1-, 5-, and 10-years respectively, lower in men compared to women. Modest improvements in survival were observed over the study period, particularly for 60-69 year old patients.

**Conclusion:** The overall prevalence of CRC in the UK has tripled in the last 20 years, leading to increased healthcare resource needs and with slight survival improvements. A worrying increasing trend of early-onset CRC is observed, warranting further research into its diagnosis and management.

## INTRODUCTION

Colorectal cancers (CRC) are the third most common cancer (more than 1.9 million cases) and the second most common cause of cancer-related death (935,000 deaths) for both men and women worldwide in 2020 ^1^. From 2016–2018 there were around 42,900 new CRC cases in the United Kingdom (UK) every year, being the fourth most common cancer in this country^2^.

The majority of CRC are sporadic and follow the adenoma-carcinoma sequence over a period of 10-15 years ^3^. There are several well-characterized risk factors such as age, family history and hereditary cancer syndromes, inflammatory bowel diseases, metabolic syndrome, sedentary lifestyle, diet, heavy alcohol consumption and smoking ^3^.

The risk of developing CRC increases with advancing age where 90% of global cases and deaths occur from age 50 years ^3^, although there has been an increase in the number of diagnoses in those under 50 years of age ^4^. CRC is more common in males ^3^ and reasons for these sex differences are potentially due to males having a higher vulnerability to environmental risk factors and higher exposure to them as well as lower uptake of CRC screening based on faecal samples ^3^. Additionally, males do not benefit from the potential protective effect of endogenous oestrogen, which has been linked to a reduction in risk in females ^5,6^.

Considering the risk factors above, there are opportunities for intervention to prevent CRC onset (primary prevention), achieve early detection (secondary prevention), or mitigate its impact on prognosis (tertiary prevention), thereby rendering it a potentially preventable disease ^7^. CRC screening has been demonstrated to be an effective and cost-effective approach ^3^ which has been implemented by numerous public health organizations worldwide including the UK ^8^. These screening programmes aim to reduce the incidence of CRC by diagnosing colorectal adenomas before CRC onset and improving prognosis by earlier detection ^7^. In the UK, the NHS (National Health Service) Bowel Cancer Screening Programme began in 2006 ^8^ and is offered every 2 years to those aged 60 to 74 with the screening now being offered from 50 years of age ^9^. The effectiveness of CRC screening has been reflected in improvements in overall survival where in England, the 5-year survival in those diagnosed through the screening programme is 81.9% but drops to 53.0% in non-screen-detected cases^10^.

Other population-based studies using cancer registries have shown that overall incidence in UK has increased but the trends indicate IRs have declined in recent years, especially after 2010, with similar trends in both sexes ^11,12^. These studies also showed an increase in the incidence of CRC in people younger than 50 years and older than 75 years of age ^11–13^. Furthermore, evidence suggests that CRC mortality in the UK has declined between 2000 to 2017 ^13^. However, despite this, the UK has one of the poorest survival rates compared to the rest of Europe and other high-income countries worldwide ^14,15^.

Understanding trends in the incidence, prevalence, and survival of CRC is an important aspect to assess the efficacy of cancer diagnosis and management to inform the development of effective preventive and screening strategies. Due to this and changes in screening age, tobacco and alcohol usage and increases of CRC in younger adults in recent years, a comprehensive assessment of the trends of CRC in relation to different population strata is lacking in the UK. Therefore, the aim of this study is to describe the population-based burden of CRC and secular trends in terms of incidence, prevalence, and survival from 2000-2021 using two primary care databases from the UK.

## METHODS

### Study design, setting, and data sources

We carried out a population cohort study using routinely collected primary care data from the UK. People with a diagnosis of CRC and a background cohort (denominator population) were identified from Clinical Practice Research Datalink (CPRD) GOLD database (July 2022) to estimate overall survival, incidence, and prevalence. We additionally carried out this study using CPRD Aurum (June 2021) to compare the results obtained from GOLD. Both these databases contain pseudonymised patient-level information on demographics, lifestyle data, clinical diagnoses, prescriptions, and preventive care provided to patients and collected by the NHS (National Health Service) as part of their care and support. CPRD GOLD contains data from across the UK whereas Aurum only contains data from England. Both databases are established primary care databases covering together over 50 million people and both were mapped to the Observational Medical Outcomes Partnership (OMOP) Common Data Model (CDM) ^16^.

### Study participants and time at risk

For the incidence and prevalence analysis, the study cohort consisted of individuals present in the database from 1st January 2000. All participants were required to be 18 years or older and have at least one year of prior history. For CPRD GOLD, these individuals were followed up to whichever came first: the cancer outcome of interest, exit from the database, date of death, or the 31st of December 2021 (the end of study period) whereas for Aurum, the end of the study period was 31st of December 2019. We studied until 2019 in CPRD Aurum as there were problems with data quality of this database post 2019 that were later resolved after this study was completed.

For the survival analysis, only individuals with a newly diagnosed cancer were included. These individuals were followed up from the date of their diagnosis to either date of death, practice stopped contributing to the database, participant left the practice or end of the study period. Any patients whose death and cancer diagnosis occurred on the same date were excluded from the survival analysis.

### Colorectal cancer definition

We used Systematized Nomenclature of Medicine Clinical Terms (SNOMED CT) diagnostic codes to identify primary CRC events. Specific diagnostics codes for malignancies of the colon and/or the rectum were included as well as unspecific codes covering these locations as these were assumed to be most likely to be from the colon or rectum. Diagnostic codes indicative of non-malignant cancer, melanoma and lymphoma were excluded as well as codes related to cancers of the appendix. For incidence and survival analyses, only diagnostic codes defining incidence disease were included whereas for prevalence estimates diagnostics codes indicative of recurrence or metastatic status of the disease were also included. The CRC cancer definition was reviewed with the aid of the CohortDiagnostics R package ^17^. This package was used to identify additional codes of interest and to remove those highlighted as irrelevant based on feedback from clinicians with oncology, primary care, and real-world data expertise through an iterative process during the initial stages of analyses. The clinical code lists used to define CRC can be found in the supplementary information S1. For survival analysis, mortality was defined as all-cause mortality based on records of date of death. OMOP-based computable phenotypes are available, together with all analytical code in

GitHub to enable research reproducibility (https://github.com/oxford-pharmacoepi/EHDENCancerIncidencePrevalence)

### Statistical methods

The population characteristics of patients with a diagnosis of CRC were summarised, with median and interquartile range (IQR) used for continuous variables, while counts and percentages were used for categorical variables.

We calculated the crude overall and annualised incidence rates (IR) and period prevalence (PP) for CRC from 2000 to 2021 in GOLD. For incidence, the number of events, the observed time at risk, and the incidence rate per 100,000 person years were summarised along with 95% confidence intervals (95% CI). Annualised IRs were calculated as the number of incident CRC cases as the numerator and the actual and exact recorded number of person-years (pys) in the general population within that year as the denominator whereas overall incidence was calculated from 2000 to 2021.

Age-standardized IRs were calculated using the 2013 European Standard Population (ESP2013) ^18^. The ESP2013 serves as a population standard with a predefined age distribution to account for differences in age structures between different populations. The ESP2013 provides predefined age distribution in five-year age bands; therefore, we collapsed these to 10-year age bands. We used the age distribution of 20-29 years from ESP2013 for age-standardization as age distributions were not available for the 18-29 years age band used in this study.

Period prevalence was calculated on 1st January for the years 2000 to 2021, with the number of patients fulfilling the case definition for CRC as the numerator. Anyone with a diagnosis of CRC was included and followed until the end of their observation period. We estimated total prevalence as although many of these cases may be considered cured after five/ten years and no longer being actively treated, people in this survivorship phase may have long-term medical needs, and therefore we considered it is important to provide accurate counts to allow for healthcare planning. The denominator was the number of participants on the 1st of January in the respective years for each database. The number of events, and prevalence (%) were summarised along with 95% confidence intervals.

For survival analysis, we used the Kaplan-Meier (KM) method to estimate the overall survival probability from observed survival times with 95% confidence intervals. We estimated the median survival and survival probability 1, 5, and 10 years after diagnosis. All results were stratified by database, by age (10-year age bands apart from the first and last age bands which were 18 to 29 years and 90 years and older respectively) and sex. Additionally, for CPRD GOLD, we stratified by calendar time of cancer diagnosis (2000-2004, 2005-2009, 2010-2014, 2015-2019 and 2020-2021). To avoid possible patient identification, we do not report results with fewer than five cases.

For Aurum, the same statistical analyses were performed using data from 1st January 2000 to 31st December 2019 to compare the results obtained from GOLD.

The statistical software R version 4.2.3 was used for analyses. For calculating incidence and prevalence, we used the IncidencePrevalence R package ^19^. For survival analysis we used the survival R package ^20^.

## RESULTS

### Patient Populations and characteristics

Overall, there were 24,340,860 and 11,388,117 eligible people available from CPRD Aurum and GOLD respectively. Attrition tables can be found in the supplementary information (Supplement S2). A summary of patient characteristics of those with a diagnosis of CRC for both databases is shown in Table 1.

**Table 1:** Baseline characteristics of colorectal cancer patients at the time of diagnosis for each database.

| Database | CPRD Aurum | CPRD GOLD |
| --- | --- | --- |
| **Number of CRC patients** | 99321 | 53797 |
| **Sex: Male (N[%])** | 55510 (55.9%) | 29800 (55.4%) |
| **Age (years) (Median [IQR])** | 72 (63 to 80) | 72 (63 to 80) |
| **Age Groups N (years) (%)** |  |  |
| 18-29 | 231 (0.2%) | 123 (0.2%) |
| 30-39 | 1182 (1.2%) | 524 (1.0%) |
| 40-49 | 3727 (3.8%) | 2001 (3.7%) |
| 50-59 | 11432 (11.5%) | 6367 (11.8%) |
| 60-69 | 24134 (24.3%) | 13466 (25.0%) |
| 70-79 | 31736 (32.0%) | 17286 (32.1%) |
| 80-89 | 22928 (23.1%) | 12118 (22.5%) |
| 90+ | 3951 (4.0%) | 1912 (3.6%) |
| **Prior history, days** |  |  |
| median [IQR] | 6623 (3,363 to 11,038) | 3493 (1,892 to 5,250) |
| **General conditions (any time prior)** |  |  |
| Atrial fibrillation | 8023 (8.1%) | 3747 (7.0%) |
| Cerebrovascular disease | 6730 (6.8%) | 3288 (6.1%) |
| Chronic liver disease | 417 (0.4%) | 201 (0.4%) |
| Chronic obstructive lung disease | 6775 (6.8%) | 3365 (6.3%) |
| Coronary arteriosclerosis | 1133 (1.1%) | 547 (1.0%) |
| Dementia | 1526 (1.5%) | 774 (1.4%) |
| Depressive disorder | 10547 (10.6%) | 5352 (9.9%) |
| Diabetes mellitus | 14553 (14.7%) | 6489 (12.1%) |
| Gastrointestinal haemorrhage | 19013 (19.1%) | 10245 (19.0%) |
| Heart failure | 3672 (3.7%) | 1796 (3.3%) |
| Hyperlipidemia | 10411 (10.5%) | 5009 (9.3%) |
| Hypertensive disorder | 40764 (41.0%) | 14932 (27.8%) |
| Ischemic heart disease | 12417 (12.5%) | 5495 (10.2%) |
| Osteoarthritis | 25022 (25.2%) | 10583 (19.7%) |
| Peripheral vascular disease | 2051 (2.1%) | 1017 (1.9%) |
| Pulmonary embolism | 1699 (1.7%) | 724 (1.3%) |
| Renal impairment | 13117 (13.2%) | 6891 (12.8%) |
| Venous thrombosis | 5749 (5.8%) | 2848 (5.3%) |

IQR: interquartile range

Overall, those with CRC are more likely to be male (∼56%), with a median age of 72 years old across both databases. The highest percentage of patients were those aged 70-79 years old, contributing to 32% of diagnosed patients.

### Incidence rates stratified by calendar year, age and sex

The overall crude IR of CRC from 2000 to 2021 was 61.5 (95% CI 61.0–62.0) per 100000 person-years for GOLD with a slightly lower IR for Aurum (56.6 95%CI 56.2 to 56.9). For GOLD, sex-specific IRs for males and females were 69.3 (95% CI 68.5–70.1) and 54.0 (95% CI 53.3–54.7) per 100000 person-years respectively, with similar results obtained for Aurum.

Annualised IRs increased across the study period for the whole population and both sexes with males having higher IRs than females (Figure 1). For males, IRs declined from 2011 to 2014-5 but increased up to 2019 for both databases. Whereas for females, in GOLD, IRs peaked in 2011 before dropping until 2019, while in Aurum IRs remained stable with a gradual increase towards the end of the study period. For GOLD there was a drop in IR in 2020 but an increase in IR in 2021 across the whole population and sex. Age standardized results using the European Standard Population for 2013 for GOLD for both sexes can be found in the supplement (Supplement S3). All results for this study can be found and downloaded in a user-friendly interactive web application: https://dpa-pde-oxford.shinyapps.io/ColorectalCancerIncPrevSurvShiny/.

**Figure 1:**
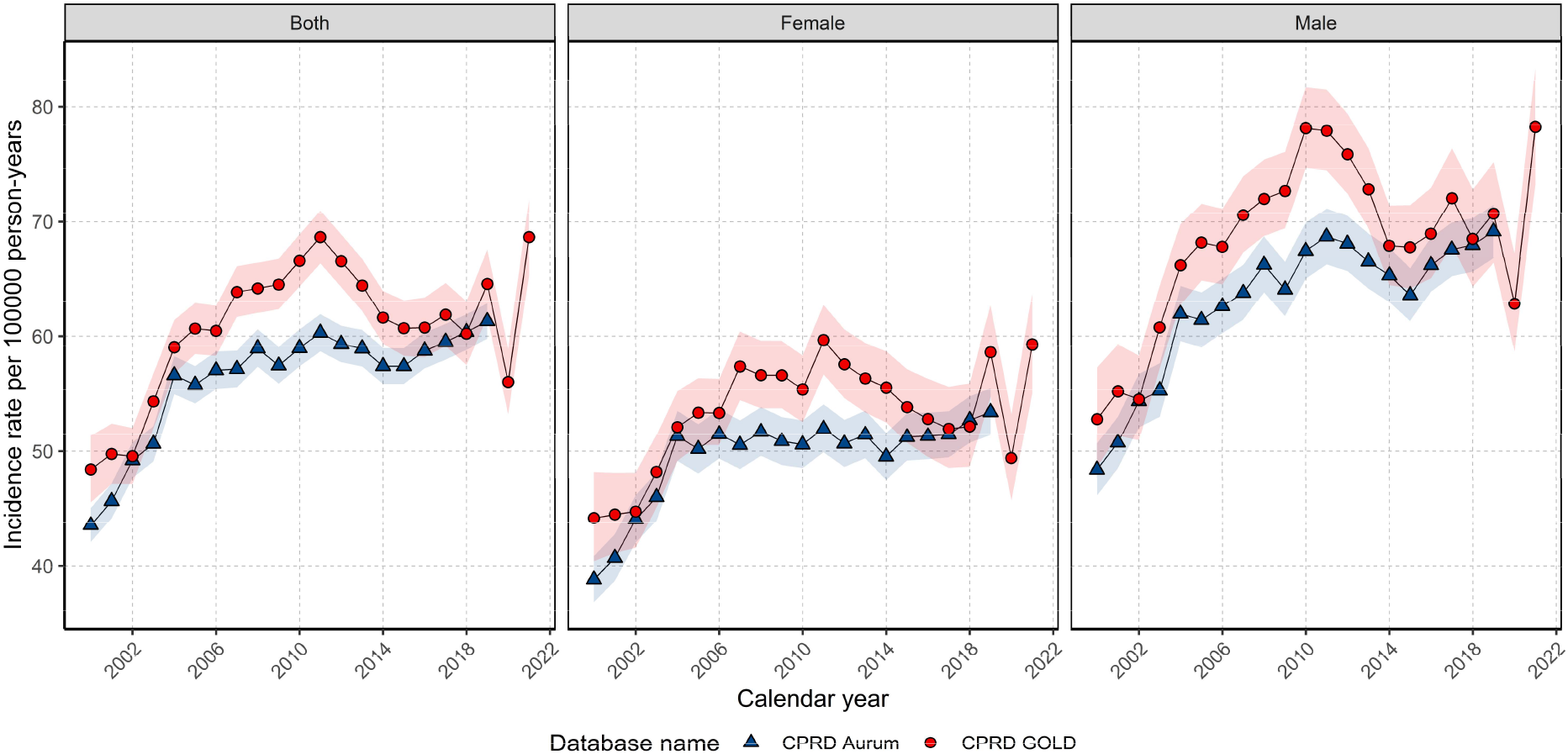
Crude annualised incidence rates for CRC from 2000 to 2021 (GOLD) and 2000-2019 (Aurum) stratified by database and sex.

Overall IRs increased with age up to 80–89 years. Those aged 18 to 29 had the lowest overall IRs across the study period with IRs of 0.77 (95% CI 0.64 to 0.92) per 100,000 person years in GOLD, whereas those aged 80–89 years-old had the highest with IRs of 277.2 (95% CI 272.3-282.1). Similar results were obtained in Aurum (Supplement S4).

Annualised IRs for each age group (Figure 2) show IRs have increased annually in those aged 30 to 59 across both databases. For those aged 60-79 years old, IRs rose and peaked in 2011 before falling and remaining stable until the end of the study period. IRs for those aged 80-89 years old remained stable after 2006. For those aged 90 and older IRs in GOLD did not increase from 2011 to the end of the study period, however in Aurum, IRs increased gradually over the study period. Across most age groups in GOLD, IRs in 2020 decreased before increasing in 2021. Stratification on both sex and age group showed similar trends in Figure 2 for both sexes. However, males had higher incidence across the study period apart from those younger than 50 years of age where there were no sex differences (Supplement S5).

**Figure 2:**
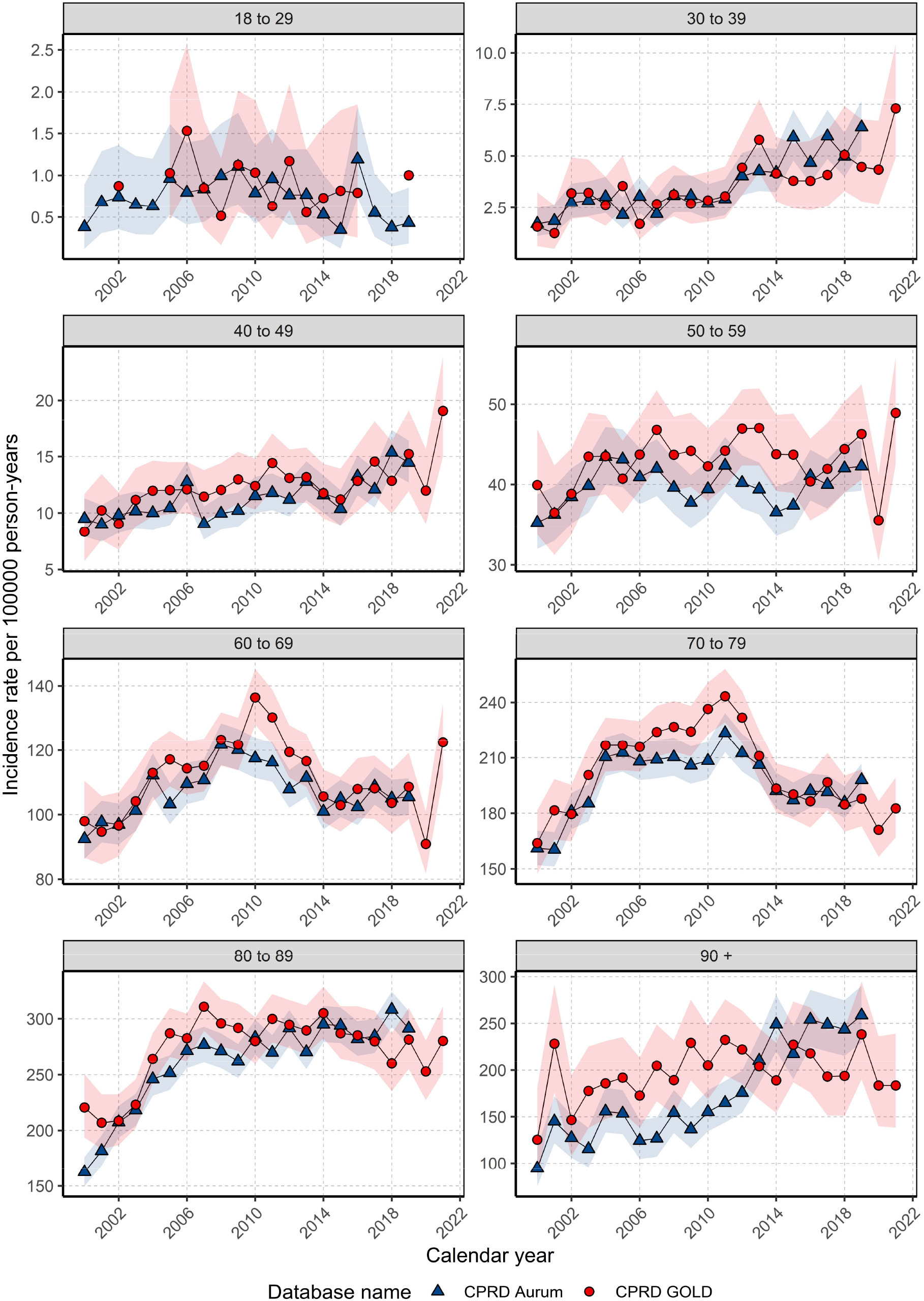
Annualised incidence rates from 2000 to 2021 stratified by database and age group.

### Prevalence for study population with database, age, and sex stratifications

In GOLD, PP for CRC in 2021 was 0.46% (0.46% to 0.47%) with similar PP in 2019 when comparing GOLD and Aurum (0.44% GOLD, 0.42% Aurum). PP in 2021 was 1.23-fold higher for males compared to females in GOLD (Males 0.51%, Females 0.42%). Annualised PP increased over the study period (Figure 3) with PP increasing 1.91-fold (Aurum) and 2.97-fold (GOLD) for the whole population, with similar increases for both females and males (Figure 3).

**Figure 3:**
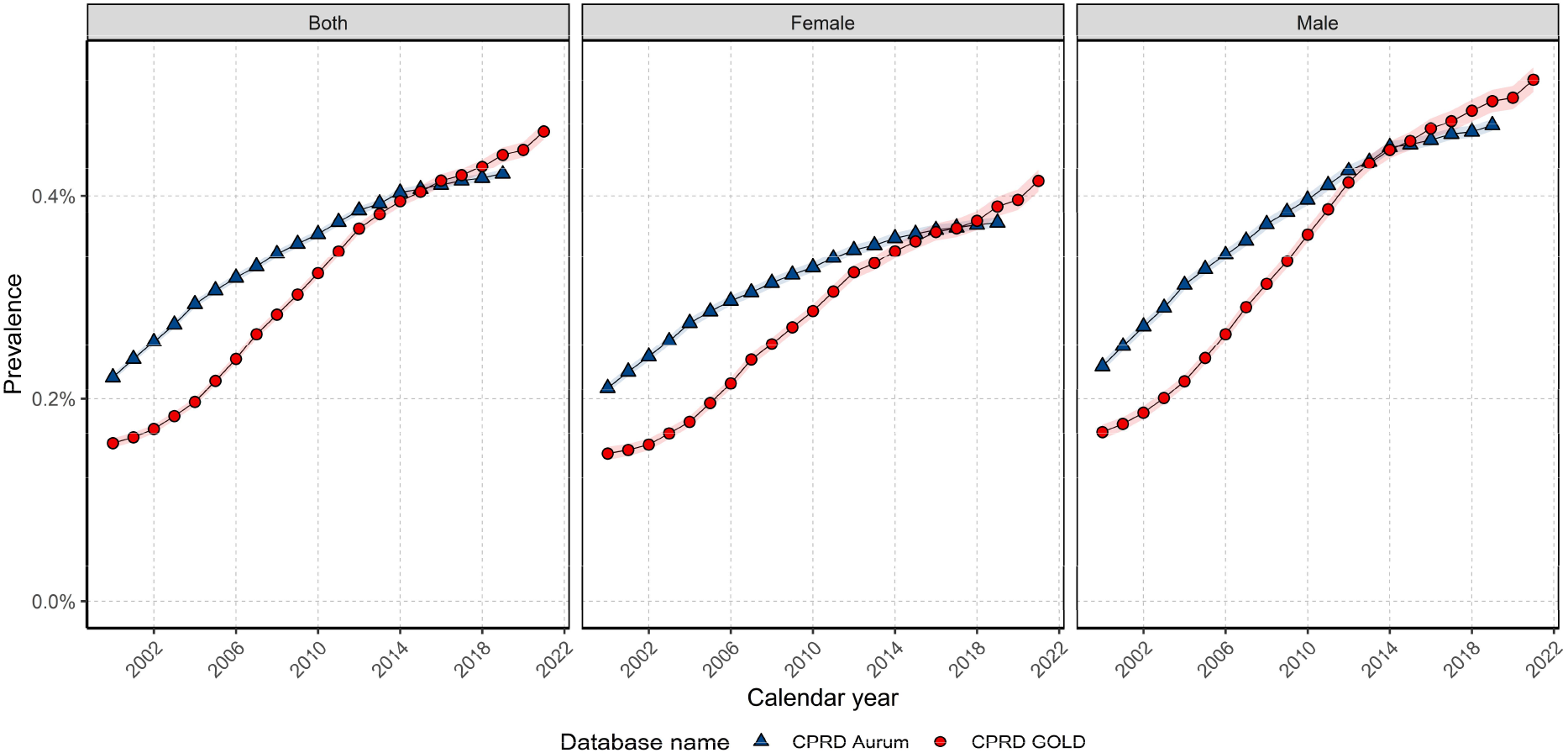
Annualised prevalence from 2000 to 2021 for whole population and stratified by sex.

When stratifying by age group, PP was highest with increasing age and was highest in those aged 80 years and older (Figure 4). PP increased then plateaued from 2011-12 for those aged between 60 to 79 years old whereas PP increased until 2007 with a small gradual decline towards the end of the study period in those aged 18 to 29 years old. Those aged 30-49 years of age and over 80 years of age showed an increase in PP across the whole study period across both databases. Stratification on both sex and age group showed similar trends to Figure 4 however males had higher prevalence across the study period compared to females apart from those younger than 50 years of age where there were no sex differences (Supplement S6).

**Figure 4:**
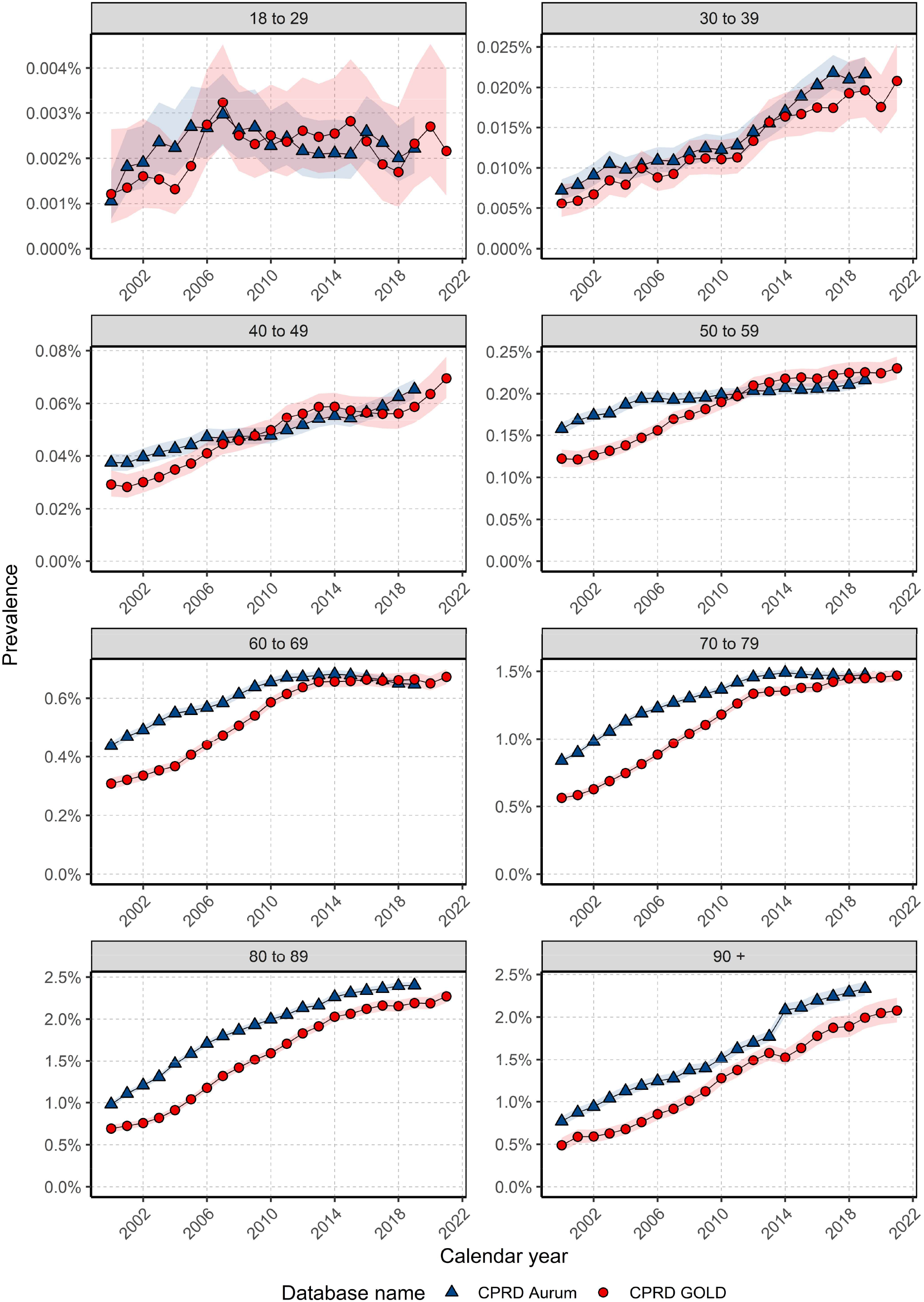
Annual prevalence from 2000 to 2021 stratified by database and age group.

### Overall survival with stratification by sex, age and calendar year

In GOLD, there were 53,098 patients with 26,084 deaths (49.1%) over the study period whereas for Aurum, there were 98,569 patients with 48,894 deaths (49.6%). Both databases had similar median follow-up time of 2.4-2.5 years and median survival of 5.44-5.54 years.

For GOLD, survival after one, five and ten years from diagnosis was 78.3%, 51.4% and 38.5% for the whole population. Females had higher median survival compared to males (Supplement S7) with median survival times for females of 6.0 (5.8 to 6.3) years and 5.1 (4.9 to 5.3) years for males with similar results obtained in Aurum (males 5.2 years; females 6.13 years). Regarding short- and long-term survival, both sexes had similar one year survival of around 79%, whereas males had lower survival at five and ten years compared to females with survival of 50.3% and 52.8% at five years and 36.1% and 40.7% at ten years for males and females respectively. Similar results were also obtained in Aurum (Supplement S8).

When stratifying by age group, median survival decreased with age for both databases. Median survival decreased in GOLD from 15.6 years in those aged 50-59 years to 0.96 years for those over 90 years (Supplement S9). Survival at one, five, and ten years, generally decreased with increasing age from 50 years and older with the lowest survival observed in those aged 90 years and older (Supplement S10). Short term survival (one year) was between 86-90% for those aged 18-49 years whereas was 49-51% in those aged 90 years and older. Long term survival at five years decreased from 50 years of age and survival at ten-year decreased with increasing age from 30 years of age across both databases.

To investigate if survival rates have changed over time, we stratified by calendar time of cancer diagnosis in five-year age windows for GOLD. Figure 5 shows the KM survival curve stratified by sex and calendar year. In GOLD one- and five-year survival slightly improved in those diagnosed in 2000 to 2004 to those diagnosed in 2015 to 2019, increasing from 77.1% (76.2-78.0) to 78.9% (78.0-79.67) for one year survival and from 48.9% (47.8-50.0) to 52.0% (50.9-53.2) for five-year survival, respectively. Stratification by sex showed improvements in five-year survival over calendar time for both sexes, but not for one year survival (Supplement S11). Stratification by different age groups showed only in those aged 60-69 years showed improvements in both one-year (80.7% [79.0-82.5] to 85.8% [84.4-97.2]) and five-year survival (55.5% [53.3-57.8] to 64.1% [61.9-66.4]) comparing those diagnosed in 2000-2004 with those diagnosed in 2015-2019 with similar results for both males and females. There was no difference in survival for those diagnosed in 2020-2021 to previous calendar year groups.

**Figure 5:**
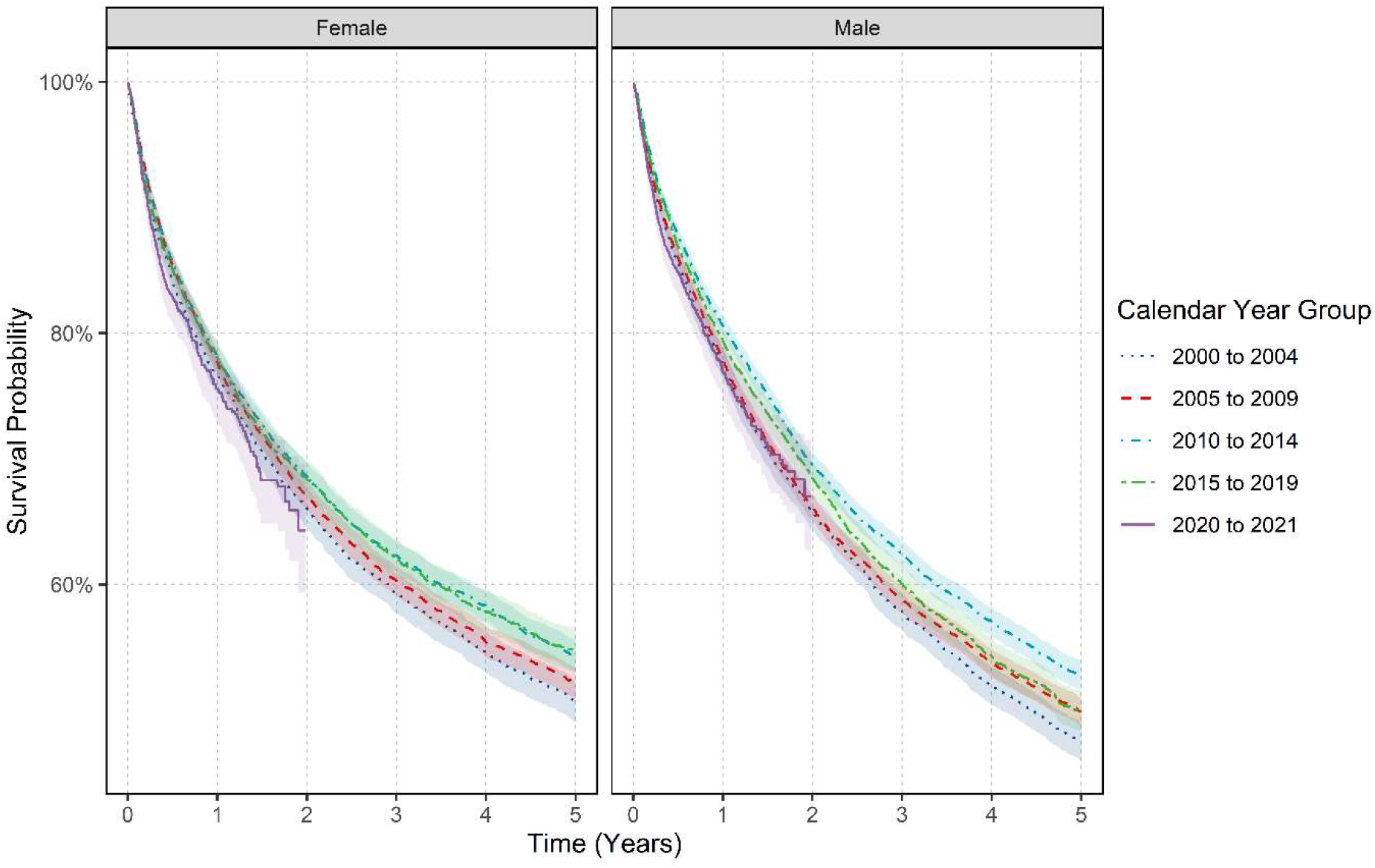
Kaplan-Meier survival curve of colorectal cancer stratified by sex and calendar year of diagnosis.

## DISCUSSION

This paper provides a comprehensive study of the trends of CRC incidence, prevalence, and survival in the UK. Incidence increased until 2011 and prevalence steadily increased over the study period, with a relevant increase in those younger than 50 years of age. Survival slightly improved from 2000 to 2019 particularly in those aged 60 to 69 years.

Our estimates of incidence rates of CRC are in line with national statistics^2^. Similar to previous literature, we also observed higher rates in men and older people ^3,21,22^. Prevalence also increased over the study period, with age group differences, likely due to the impact of screening programmes, as we observed a stabilisation during the last decade of the study in the targeted age groups ^23^.

We observed four main trends on incidence during the study period. First, a progressive increase after 2006, reaching its peak in 2011 and decreasing the subsequent years, with a sharp rise in 2021, likely driven by deferred diagnosis after the pandemic ^24^. Earlier trends could be associated with the introduction of the National Bowel Screening Programme in 2006^25^. The detection of early-stage asymptomatic cases could explain an increase in IRs with a subsequent reduction due to endoscopic resection of adenomas ^7,26^. This pattern had already been described previously ^12,13^. A population-based study in the Netherlands showed an initial increase in incidence after the start of the screening programme, especially in early-stage CRC incidence, followed by a decrease in both early- and advanced-stage CRC incidence ^27^.

Third, we observed a worrying increase in cases in those under 50 years of age, which has also been observed in many other countries ^4,12^. The reasons for this increase in early-onset colorectal cancer (EOCRC) are unclear. EOCRC appears to have differential clinical and pathological characteristics^28^. One possible explanation is birth cohort effects ^29,30^, where those born after 1960’s were exposed to modifiable risk factors such as unhealthy Western diets, sedentary behaviour, and lower physical activity ^29,31^. Interestingly, we did not observe sex differences in this age group, suggesting these exposures are not sex specific ^3,30,32^. On the other hand, the rising incidence of CRC in younger age groups has led to some countries, including the UK, to lower the bowel cancer screening age eligibility ^9^. Notably, a decline in young-onset CRC is observed in those countries with a lowered age for CRC screening eligibility (Austria and Italy), while an increasing incidence is observed in other nine high-income countries (Australia, Canada, Denmark, Germany, New Zealand, Slovenia, Sweden, UK and USA) ^30^.

Fourth, the IRs also increased over the study period in nonagenarians. This could be due to improvements in life expectancy ^33^. Also, this age group does not benefit of routine cancer screening, as it is not recommended over 85 years of age ^33^. The diagnostic and treatment in this age group is challenging, as colonoscopy is an invasive procedure associated with increased risks and incomplete examinations in the elderly or frail patients ^34^. On the other hand, several studies point out that the age itself should not be a relevant criterion and suggest geriatric assessment to optimise cancer care pathways as well as including older patients in clinical trials ^35,36^. Notwithstanding the above, a north English population-based study found that age remains a major factor in treatment decisions in the UK ^37^.

Sex differences for CRC incidence and survival have previously been described ^3,6,22,38^. We observed higher IRs and worse overall survival in males, except for EOCRC. Negligible differences in age-standardized survival among men and women had been previously described in England ^22^, while other studies had reported better overall and cancer-specific survival in women than men as well as better recurrence-free survival ^39^. These sex differences can be explained by several modifiable and non-modifiable factors. Males have higher exposure to environmental risk factors (obesity, unhealthy diet, and alcohol consumption), and higher vulnerability to them ^3,6,40^. Men also have lower uptake of CRC screening, although in England a higher proportion of CRCs in males are diagnosed via the screening programme ^22^. This could be explained by the fact that women are more prone to have proximal and more aggressive cancers and because gFOBT/FIT screening is less effective in women ^22^. On the other hand, sexual dimorphisms also could contribute to these differences ^6^.

Short- and long-term survival results are in line with national statistics ^41^ and with previous literature ^14^. Median survival decreased with age as expected, which can be explained by frailty and comorbidities ^42^.

Despite the improvement in therapeutic management in recent years, we only observed a modest impact on survival. Moreover, when stratifying by age, only those aged 60-69 years had consistent improvements, which could reflect the effects of the screening in this age group^10^. Survival rates in the UK have already been assessed in other studies. Compared to other European countries with similar health systems and populations, overall survival rates are worse in the UK, particularly for elderly people ^14,15^ Careful consideration is necessary when interpreting these comparisons, since these studies are based on cancer registries and there are intercountry differences in data collection, potentially introducing bias ^43^. Greater efforts are being made to understand the extensive heterogeneity of CRC to avoid the current “one-fits-for-all” therapeutic strategy, with emerging research moving closer to personalized treatments based on the molecular characteristics of the tumour and enhancing risk stratification, chemotherapy monitoring, and early relapse detection ^44,45^. Once this research can be applied in clinical practice, it is expected that survival rates will enhance in the coming years.

This study has many strengths. First, we used two large primary care databases covering a large proportion of the UK, including England and the devolved nations ^46,47^. The similarity between the results from both databases proves the robustness of our findings. The sharp increase in incidence at the start of the study period between 2000-2004 could be explained by the introduction of the Quality and Outcomes Framework (QOF) in 2004 which encouraged general practitioners to record all new cases ^48^. Second, our study used a complete representative population-based database for the assessment of both numerators and denominators in the estimated incidence and prevalence. In contrast, cancer registry studies extrapolate the registry data to the whole population using national statistics which could potentially introduce bias ^49,50^. Third, the high validity of mortality data with over 98% accuracy^51^ allowed us to examine the impact of calendar time on overall survival.

Our study also had limitations. Firstly, we used primary care data without linkage to cancer registry, potentially leading to misclassification and delayed recording of diagnoses. However, previous validation studies have shown high accuracy and completeness of cancer diagnoses in UK primary care records ^52^. Secondly, our use of primary care records precluded us from studying tumour histology, genetic mutations, staging or cancer therapies which can all impact survival. Therefore, our survival estimates may overestimate survival in those with higher staging ^10^ as well those with specific genetic mutations such as KRAS and BRAF ^53,54^. Other factors such as socio-economic status and ethnicity could also result in different values for incidence, prevalence, and survival ^55^.

## CONCLUSION

In conclusion, our study shows a reduction in CRC incidence over the years mostly driven by those aged 60-79 years, in whom consistent small improvements in survival are also observed, which could be associated with screening. All this leads to an overall increase in CRC prevalence, which will require increased healthcare resources. On the other hand, a worryingly increase in the incidence of EOCRC raises the question whether lowering the screening age could contribute to better outcomes in these patients. It also reflects the necessity of a better understanding of the biology of young-onset CRC and associated risk factors, which will facilitate and expand risk-stratified screening in the future. Finally, despite advancements in therapeutic management, we only found modest improvements in survival which can be explained by the convergence of multiple factors and underscores the need for further research into the management of CRC.

## ADDITIONAL INFORMATION

## Data Availability

Patient level data used in this study was obtained through an approved application to the CPRD (application number 22_001843) and is only available following an approval process to safeguard the confidentiality of patient data. Details on how to apply for data access can be found at https://cprd.com/data-access.

https://github.com/oxford-pharmacoepi/EHDENCancerIncidencePrevalence

https://dpa-pde-oxford.shinyapps.io/ColorectalCancerIncPrevSurvShiny/

## ACKNOWLEDGEMENTS

None.

## AUTHOR CONTRIBUTIONS

All authors were involved in the study conception and design, interpretation of the results, and the preparation of the manuscript. AD and WYM implemented the data curation, data harmonisation, data quality tests and assessment. DN carried out data analysis for the manuscript. AWR developed the clinical code lists for CRC for this study. AG and AWR reviewed the clinical codelists used in this study. PPP, CGA and DN wrote the initial draft of the manuscript with DPA. DN, EB, AD, WYM and DPA had access to the CPRD data. All authors critically reviewed the final manuscript and gave consent for publication.

## ETHICS APPROVAL AND CONSENT TO PARTICIPATE

The use of CPRD data for this study was approved by the Independent Scientific Advisory Committee (22_001843). Informed consent was waived by competent authorities due to the anonymized nature of patient data.

## CONSENT FOR PUBLICATION

Not applicable.

## DATA AVAILABILITY

This study is based in part on data from the Clinical Practice Research Datalink (CPRD) obtained under licence from the UK Medicines and Healthcare products Regulatory Agency. The data is provided by patients and collected by the NHS as part of their care and support. The interpretation and conclusions contained in this study are those of the author/s alone. Patient level data used in this study was obtained through an approved application to the CPRD (application number 22_001843) and is only available following an approval process to safeguard the confidentiality of patient data. Details on how to apply for data access can be found at https://cprd.com/data-access.

## ABBREVIATIONS

CPRD: Clinical Practice Research Datalink
CRC: colorectal cancer
EOCRC: early-onset colorectal cancer
FIT: Faecal Immunochemical Test
gFOBT: guaiac Faecal Occult Blood Test
IR: incidence rate
IQR: interquartile range
KM: Kaplan-Meier
NHS: National Health Service
OS: overall survival
PP: period prevalence
Pys: person-years
SNOMED CT: Systematized Nomenclature of Medicine - Clinical Terms

