## Supplementary for "Incidence, prevalence, and survival of colorectal cancer in the United Kingdom from 2000-2021: a population-based cohort study"

### **S1: Clinical codelists for colorectal cancer**

The clinical codelists used for colorectal cancer is listed in the table below with the corresponding SNOMED concept ID, OMOP concept ID and concept description. Only diagnosis records alone were used to identify cancer outcome for this study. Different codelists were created for incident and prevalent definitions of colorectal cancer. We developed concept definitions using ATLAS, the OHDSI open-source platform (<https://github.com/OHDSI/atlas>). Clinical adjudicators reviewed the cohort definitions and associated concept sets.

| **Concept Id** | **Concept SNOMED Code** | **Concept Description** | **Code used for** |
| --- | --- | --- | --- |
| 4180790 | 363406005 | Malignant tumor of colon | Incidence and prevalence |
| 443390 | 363351006 | Malignant tumor of rectum | Incidence and prevalence |
| 4110575 | 254582000 | Adenocarcinoma of rectum | Incidence and prevalence |
| 443381 | 363410008 | Malignant tumor of sigmoid colon | Incidence and prevalence |
| 443391 | 363350007 | Malignant tumor of cecum | Incidence and prevalence |
| 435754 | 363412000 | Malignant tumor of ascending colon | Incidence and prevalence |
| 4180792 | 363414004 | Malignant tumor of rectosigmoid junction | Incidence and prevalence |
| 443384 | 363408006 | Malignant tumor of transverse colon | Incidence and prevalence |
| 4116240 | 255081007 | Carcinoma of cecum | Incidence and prevalence |
| 4180791 | 363407001 | Malignant tumor of hepatic flexure | Incidence and prevalence |
| 443382 | 363409003 | Malignant tumor of descending colon | Incidence and prevalence |
| 4181344 | 363413005 | Malignant tumor of splenic flexure | Incidence and prevalence |
| 4198567 | 315058005 | HNPCC - hereditary nonpolyposis colon cancer | Incidence and prevalence |
| 4089661 | 187757001 | Malignant neoplasm, overlapping lesion of colon | Incidence and prevalence |
| 74582 | 93984006 | Primary malignant neoplasm of rectum | Incidence and prevalence |
| 79740 | 109838007 | Overlapping malignant neoplasm of colon | Incidence and prevalence |
| 197500 | 93761005 | Primary malignant neoplasm of colon | Incidence and prevalence |
| 432257 | 94105000 | Primary malignant neoplasm of transverse colon | Incidence and prevalence |
| 432837 | 371977004 | Primary malignant neoplasm of cecum | Incidence and prevalence |
| 436635 | 94006002 | Primary malignant neoplasm of sigmoid colon | Incidence and prevalence |
| 437798 | 94072004 | Primary malignant neoplasm of splenic flexure of colon | Incidence and prevalence |
| 438090 | 109839004 | Overlapping malignant neoplasm of rectum, anus and anal canal | Incidence and prevalence |
| 438699 | 93980002 | Primary malignant neoplasm of rectosigmoid junction | Incidence and prevalence |
| 438979 | 93826009 | Primary malignant neoplasm of hepatic flexure of colon | Incidence and prevalence |
| 441800 | 93771007 | Primary malignant neoplasm of descending colon | Incidence and prevalence |
| 443396 | 363510005 | Malignant tumor of large intestine | Incidence and prevalence |
| 764981 | 98981000119103 | Primary malignant neoplasm of ileocecal valve | Incidence and prevalence |
| 4115028 | 285312008 | Carcinoma of sigmoid colon | Incidence and prevalence |
| 4149847 | 269533000 | Carcinoma of colon | Incidence and prevalence |
| 4151260 | 269544008 | Carcinoma of the rectosigmoid junction | Incidence and prevalence |
| 4184850 | 413446001 | Adenocarcinoma of cecum | Incidence and prevalence |
| 4193165 | 312113007 | Carcinoma of descending colon | Incidence and prevalence |
| 4193871 | 312112002 | Carcinoma of transverse colon | Incidence and prevalence |
| 4193872 | 312115000 | Carcinoma of splenic flexure | Incidence and prevalence |
| 4200514 | 301756000 | Adenocarcinoma of sigmoid colon | Incidence and prevalence |
| 4207182 | 312111009 | Carcinoma of ascending colon | Incidence and prevalence |
| 4207183 | 312114001 | Carcinoma of hepatic flexure | Incidence and prevalence |
| 4246125 | 93854002 | Primary malignant neoplasm of large intestine | Incidence and prevalence |
| 4247719 | 93683002 | Primary malignant neoplasm of ascending colon | Incidence and prevalence |
| 4256776 | 408645001 | Adenocarcinoma of large intestine | Incidence and prevalence |
| 4307687 | 422581008 | Carcinoma of colon, stage II | Incidence and prevalence |
| 4310858 | 422375001 | Carcinoma of colon, stage III | Incidence and prevalence |
| 4312001 | 422985007 | Carcinoma of colon, stage IV | Incidence and prevalence |
| 4312240 | 425213009 | Carcinoma of colon, stage I | Incidence and prevalence |
| 4322376 | 425178004 | Adenocarcinoma of rectosigmoid junction | Incidence and prevalence |
| 36683531 | 781382000 | Malignant neoplasm of colon and/or rectum | Incidence and prevalence |
| 36713361 | 681601000119101 | Primary adenocarcinoma of ascending colon | Incidence and prevalence |
| 36715911 | 721695008 | Primary adenocarcinoma of ascending colon and right flexure | Incidence and prevalence |
| 36715912 | 721696009 | Primary adenocarcinoma of transverse colon | Incidence and prevalence |
| 36717495 | 721699002 | Primary adenocarcinoma of descending colon and splenic flexure | Incidence and prevalence |
| 37016239 | 184881000119106 | Primary adenocarcinoma of rectosigmoid junction | Incidence and prevalence |
| 37018659 | 96281000119107 | Overlapping malignant neoplasm of colon and rectum | Incidence and prevalence |
| 37208245 | 681651000119102 | Primary adenocarcinoma of descending colon | Incidence and prevalence |
| 40492939 | 448994001 | Carcinoma of upper rectum | Incidence and prevalence |
| 42537577 | 737058005 | Microsatellite instability-high colorectal cancer | Incidence and prevalence |
| 42872396 | 1701000119104 | Primary adenocarcinoma of colon | Incidence and prevalence |
| 4196256 | 314966008 | Local recurrence of malignant tumor of rectum | Prevalence only |
| 4196264 | 314998002 | Metastasis from malignant tumor of colon | Prevalence only |

### **S2: Population attrition showing eligible patients for study from each database.**

| **N** | **Reason** | **N excluded** | **Database** |
| --- | --- | --- | --- |
| 39999011 | Starting population |  | Aurum |
| 39999011 | Missing year of birth | 0 |  |
| 39999011 | Missing sex | 0 |  |
| 34833388 | Cannot satisfy age criteria during the study period based on year of birth | 5165623 |  |
| 29190480 | No observation time available during study period | 5642908 |  |
| 29190480 | Doesn't satisfy age criteria during the study period | 0 |  |
| 25483313 | Prior history requirement not fulfilled during study period | 3707167 |  |
| 24340860 | No observation time available after applying age and prior history criteria | 1142453 |  |
| 24340860 | Starting analysis population |  |  |
| 24340860 | Estimating prevalence |  |  |
| 24319115 | Excluded due to prior event (do not pass outcome washout during study period) | 21745 |  |
| 24319115 | Estimating incidence |  |  |
| 99321 | With a cancer diagnosis | 24219794 |  |
| 98569 | Cancer diagnosis not on same date as death | 752 |  |
| 86710 | Estimating survival |  |  |
| 17054819 | Starting population |  | GOLD |
| 17054819 | Missing year of birth | 0 |  |
| 17054819 | Missing sex | 0 |  |
| 15210165 | Cannot satisfy age criteria during the study period based on year of birth | 1844654 |  |
| 13978229 | No observation time available during study period | 1231936 |  |
| 13978229 | Doesn't satisfy age criteria during the study period | 0 |  |
| 12254874 | Prior history requirement not fulfilled during study period | 1723355 |  |
| 11388117 | No observation time available after applying age and prior history criteria | 866757 |  |
| 11388117 | Starting analysis population |  |  |
| 11388117 | Estimating prevalence |  |  |
| 11381775 | Excluded due to prior event (do not pass outcome washout during study period) | 6342 |  |
| 11381775 | Estimating incidence |  |  |
| 53797 | With a cancer diagnosis | 11327978 |  |
| 53098 | Cancer diagnosis not on same date as death | 699 |  |
| 53098 | Estimating survival |  |  |

### **S3: Age standardised by the European Standard Population for incidence rates for CPRD GOLD for colorectal cancer stratified by sex.**

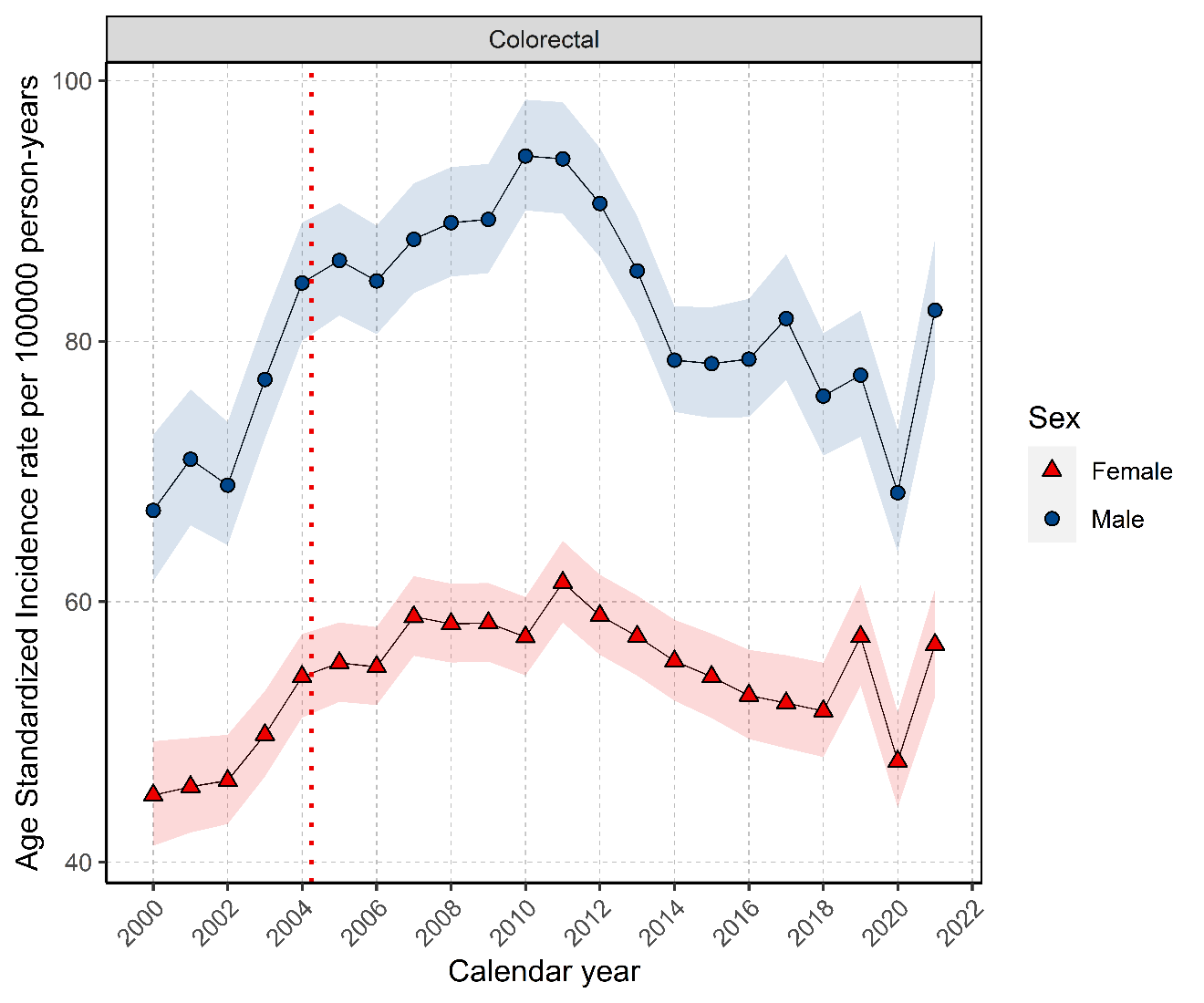

### **S4: Overall incidence rate for colorectal cancer from stratified by database and age group.**

| **Age Group** | **n persons** | **person years** | **n events** | **Incidence (100000 pys)** | **Database** |
| --- | --- | --- | --- | --- | --- |
| 18 to 29 | 9,238,374 | 31,941,017 | 231 | 0.72 (0.63 to 0.82) | CPRD Aurum |
| 30 to 39 | 8,292,235 | 32,676,728 | 1,182 | 3.62 (3.41 to 3.83) |  |
| 40 to 49 | 6,514,825 | 32,912,807 | 3,727 | 11.32 (10.96 to 11.69) |  |
| 50 to 59 | 5,434,527 | 28,632,117 | 11,432 | 39.93 (39.20 to 40.67) |  |
| 60 to 69 | 4,161,289 | 22,412,580 | 24,134 | 107.68 (106.33 to 109.05) |  |
| 70 to 79 | 3,102,304 | 16,108,479 | 31,736 | 197.01 (194.85 to 199.19) |  |
| 80 to 89 | 1,930,511 | 8,707,470 | 22,928 | 263.31 (259.92 to 266.74) |  |
| 90 + | 658,438 | 2,239,680 | 3,951 | 176.41 (170.95 to 182.00) |  |
| 18 to 29 | 3,871,109 | 16,018,361 | 123 | 0.77 (0.64 to 0.92) | CPRD GOLD |
| 30 to 39 | 3,682,270 | 15,106,025 | 524 | 3.47 (3.18 to 3.78) |  |
| 40 to 49 | 3,246,464 | 16,111,285 | 2,001 | 12.42 (11.88 to 12.98) |  |
| 50 to 59 | 2,884,106 | 14,703,972 | 6,367 | 43.30 (42.24 to 44.38) |  |
| 60 to 69 | 2,289,865 | 11,852,052 | 13,466 | 113.62 (111.71 to 115.55) |  |
| 70 to 79 | 1,675,784 | 8,346,824 | 17,286 | 207.10 (204.02 to 210.21) |  |
| 80 to 89 | 1,016,741 | 4,372,137 | 12,118 | 277.16 (272.25 to 282.14) |  |
| 90 + | 319,907 | 955,339 | 1,913 | 200.24 (191.37 to 209.42) |  |

**Pys: person years**

### **S5: Annualised incidence rates for colorectal cancer stratified by database, sex, and age group.**

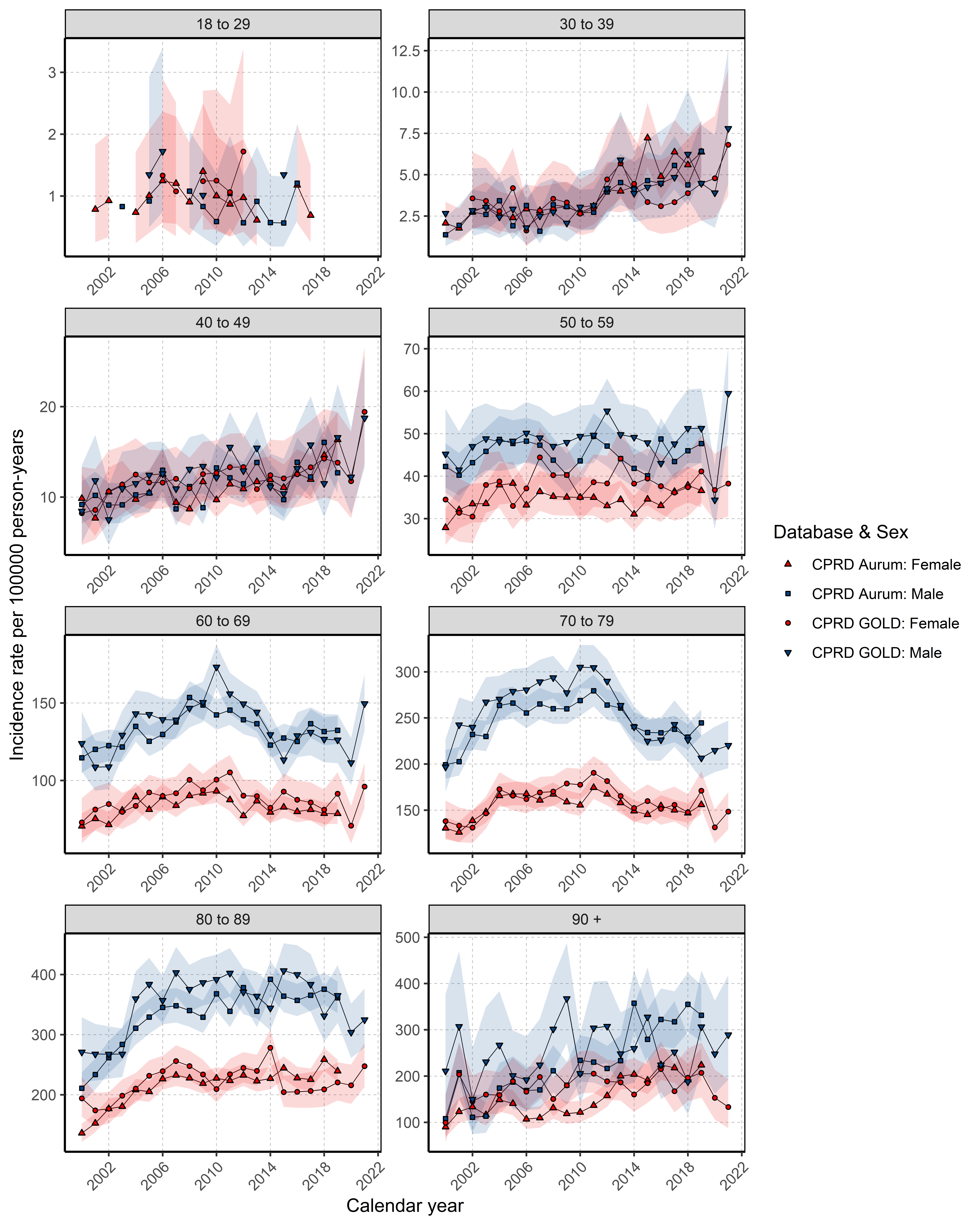

### **S6: Annualised prevalence for colorectal cancer stratified by database, sex and age group.**

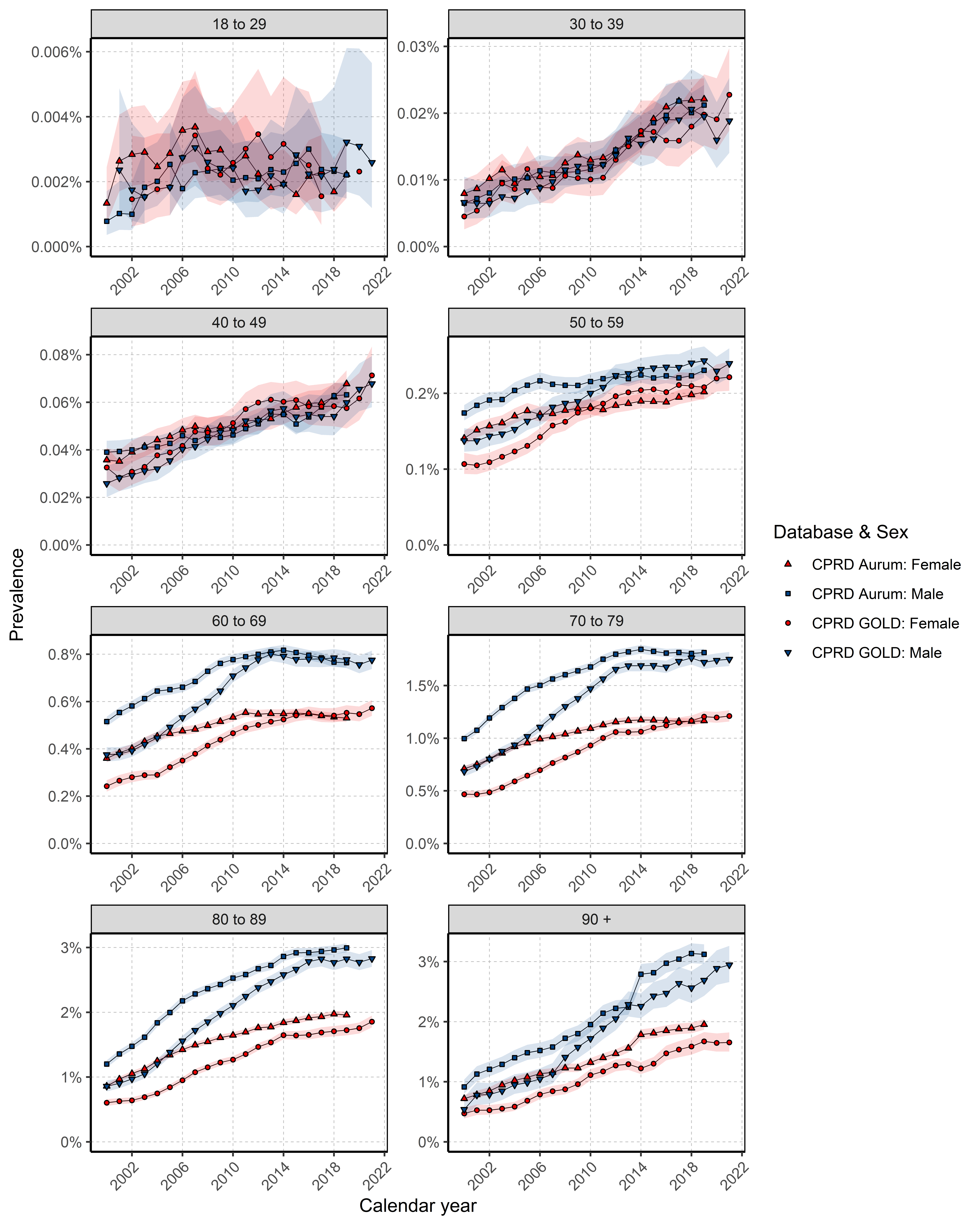

### **S7: Kaplan-Meier survival curve of colorectal cancer by database and sex**

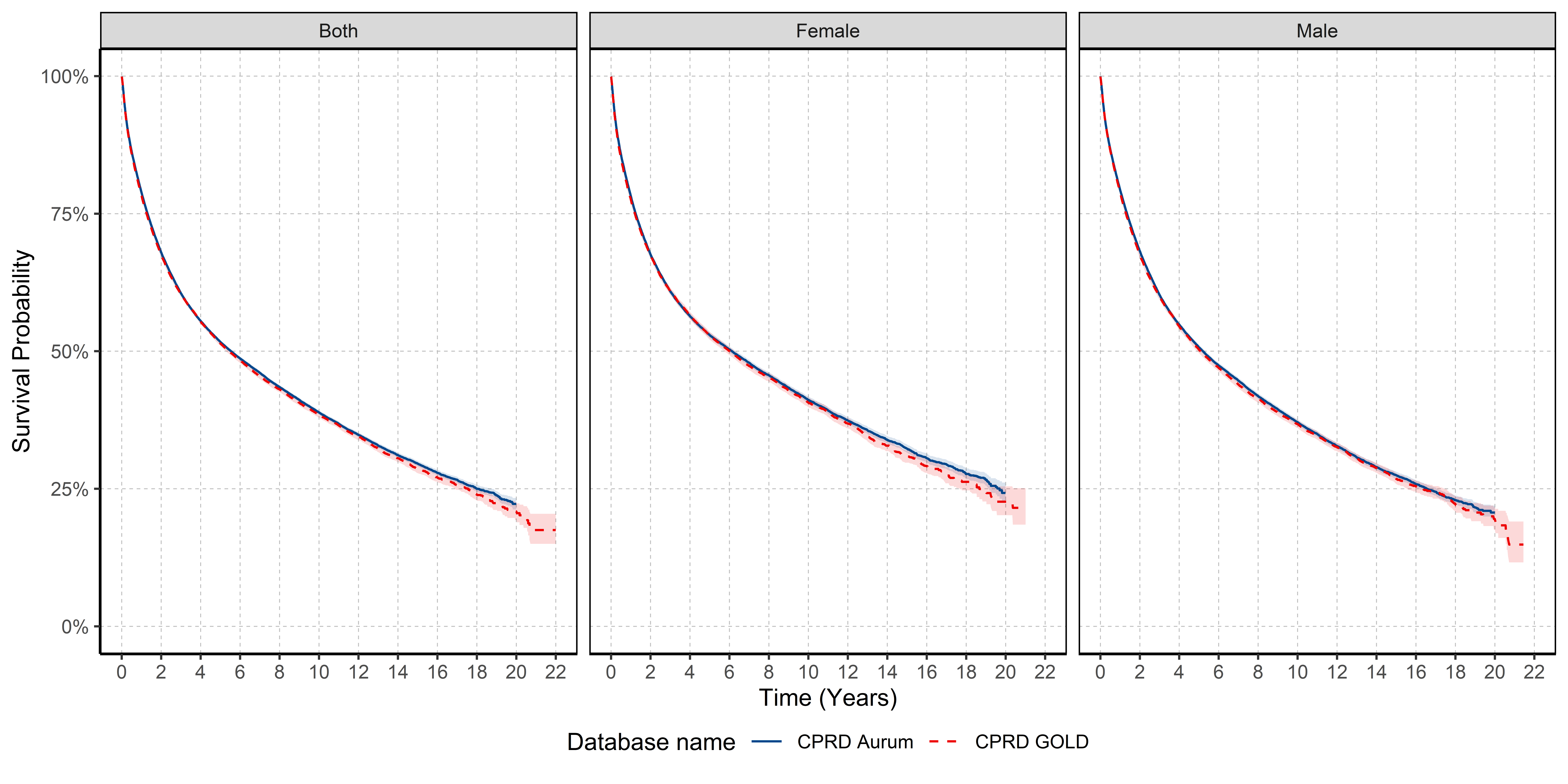

### **S8: Survival (%) after 1, 5 and 10 years after colorectal cancer diagnosis stratified by database and sex.**

| **Time** | **Sex** | **% Survival (95% CI)** | **Database** |
| --- | --- | --- | --- |
| 1 | Male | 79.4 (79.0 - 79.7) | Aurum |
| 5 |  | 50.7 (50.3 - 51.2) |  |
| 10 |  | 37.1 (36.6 - 37.6) |  |
| 1 | Female | 78.2 (77.8 - 78.6) |  |
| 5 |  | 52.9 (52.4 - 53.5) |  |
| 10 |  | 41.2 (40.6 - 41.8) |  |
| 1 | Male | 78.9 (78.4 - 79.4) | GOLD |
| 5 |  | 50.3 (49.7 – 51.0) |  |
| 10 |  | 36.8 (36.1 - 37.5) |  |
| 1 | Female | 77.6 (77.1 - 78.2) |  |
| 5 |  | 52.8 (52.1 - 53.5) |  |
| 10 |  | 40.7 (39.8 - 41.5) |  |

### **S9: Median survival stratified by database and age group.**

| **Age Group** | **Median survival in years (%95 CI)** | **n persons** | **n events** | **Database** |
| --- | --- | --- | --- | --- |
| 18 to 29 | Not achieved | 230 | 76 | Aurum |
| 30 to 39 | Not achieved | 1178 | 346 |  |
| 40 to 49 | Not achieved | 3714 | 1267 |  |
| 50 to 59 | 15.3 (13.8 - 16.4) | 11389 | 4162 |  |
| 60 to 69 | 10.8 (10.4 - 11.3) | 24043 | 9774 |  |
| 70 to 79 | 5.8 (5.6 - 6.0) | 31523 | 16368 |  |
| 80 to 89 | 2.6 (2.5 - 2.7) | 22648 | 14175 |  |
| 90 + | 1.0 (1.0 - 1.1) | 3844 | 2726 |  |
| 18 to 29 | Not achieved | 122 | 29 | GOLD |
| 30 to 39 | Not achieved | 522 | 145 |  |
| 40 to 49 | Not achieved | 1991 | 682 |  |
| 50 to 59 | 15.6 (13.8 - 19.3) | 6322 | 2227 |  |
| 60 to 69 | 10.4 (9.8 - 11.1) | 13375 | 5324 |  |
| 70 to 79 | 5.6 (5.3 - 5.8) | 17083 | 8808 |  |
| 80 to 89 | 2.5 (2.4 - 2.6) | 11860 | 7525 |  |
| 90 + | 1.0 (0.9 - 1.1) | 1823 | 1344 |  |

Not achieved: Median survival was not achieved in study period.

### **S10: One-, five- and ten-year survival (95% confidence intervals) of colorectal cancer stratified by database and age group.**

| **Age Group** | **One-year Survival (%)** | | **Five-year Survival (%)** | | **Ten-year Survival (%)** | |
| --- | --- | --- | --- | --- | --- | --- |
|  | **Aurum** | **GOLD** | **Aurum** | **GOLD** | **Aurum** | **GOLD** |
| 18-29 | 87.2 (82.8 - 91.8) | 89.6 (84.2 - 95.4) | 60.9 (53.9 - 68.8) | 70.8 (61.9 - 80.9) | 54.5 (46.8 - 63.4) | 67.4 (57.2 - 79.5) |
| 30-39 | 86.4 (84.3 - 88.5) | 89.5 (86.8 - 92.3) | 65.4 (62.3 - 68.8) | 67.1 (62.5 – 72.0) | 58.6 (54.9 - 62.5) | 63.0 (58.0 - 68.5) |
| 40-49 | 87.4 (86.3 - 88.5) | 85.5 (83.9 - 87.1) | 63.3 (61.6 - 65.1) | 62.8 (60.5 - 65.3) | 57.7 (55.8 - 59.6) | 57.8 (55.3 - 60.5) |
| 50-59 | 87.3 (86.6 - 87.9) | 86.6 (85.8 - 87.5) | 62.5 (61.5 - 63.5) | 63.7 (62.4 - 65.1) | 55.1 (54.0 - 56.2) | 55.7 (54.2 - 57.3) |
| 60-69 | 85.3 (84.9 - 85.8) | 84.7 (84.0 - 85.3) | 61.9 (61.2 - 62.6) | 61.3 (60.3 - 62.2) | 51.5 (50.7 - 52.2) | 50.7 (49.6 - 51.8) |
| 70-79 | 79.6 (79.2 - 80.1) | 79.2 (78.6 - 79.8) | 52.7 (52.1 - 53.3) | 52.1 (51.3 - 52.9) | 37.2 (36.5 - 37.9) | 36.3 (35.3 - 37.2) |
| 80-89 | 69.2 (68.5 - 69.8) | 68.1 (67.2 - 68.9) | 36.1 (35.4 - 36.8) | 34.4 (33.5 - 35.5) | 17.7 (16.9 - 18.5) | 16.8 (15.8 - 17.9) |
| 90+ | 51 (49.3 - 52.7) | 48.7 (46.4 - 51.2) | 14 (12.6 - 15.5) | 15.1 (13.1 - 17.3) | 3.3 (2.2 - 4.8) | 5.0 (3.4 - 7.4) |

### **S11: Survival after 1 and 5 years stratified by database, calendar year for whole population and sex.**

| **Calendar Year** | **Time (years)** | **% Survival (95% CI)** | **Sex** | **Database** |
| --- | --- | --- | --- | --- |
| 2000 to 2004 | 1 | 79.82 (79.21 - 80.43) | Both | CPRD Aurum |
| 2005 to 2009 | 1 | 77.85 (77.29 - 78.40) |  |  |
| 2010 to 2014 | 1 | 78.80 (78.28 - 79.32) |  |  |
| 2015 to 2019 | 1 | 79.71 (79.21 - 80.21) |  |  |
| 2000 to 2004 | 1 | 76.80 (75.82 - 77.79) |  | CPRD GOLD |
| 2005 to 2009 | 1 | 77.66 (76.96 - 78.36) |  |  |
| 2010 to 2014 | 1 | 79.54 (78.88 - 80.21) |  |  |
| 2015 to 2019 | 1 | 79.30 (78.46 - 80.14) |  |  |
| 2020 to 2021 | 1 | 76.32 (74.57 - 78.12) |  |  |
| 2000 to 2004 | 1 | 79.23 (78.32 - 80.15) | Female | CPRD Aurum |
| 2005 to 2009 | 1 | 77.53 (76.70 - 78.37) |  |  |
| 2010 to 2014 | 1 | 78.25 (77.46 - 79.06) |  |  |
| 2015 to 2019 | 1 | 78.65 (77.88 - 79.42) |  |  |
| 2000 to 2004 | 1 | 76.48 (75.02 - 77.97) |  | CPRD GOLD |
| 2005 to 2009 | 1 | 77.44 (76.40 - 78.50) |  |  |
| 2010 to 2014 | 1 | 78.11 (77.09 - 79.15) |  |  |
| 2015 to 2019 | 1 | 78.29 (77.02 - 79.58) |  |  |
| 2020 to 2021 | 1 | 75.59 (72.94 - 78.32) |  |  |
| 2000 to 2004 | 1 | 80.30 (79.49 - 81.12) | Male | CPRD Aurum |
| 2005 to 2009 | 1 | 78.10 (77.36 - 78.85) |  |  |
| 2010 to 2014 | 1 | 79.22 (78.53 - 79.90) |  |  |
| 2015 to 2019 | 1 | 80.53 (79.88 - 81.19) |  |  |
| 2000 to 2004 | 1 | 77.07 (75.75 - 78.41) |  | CPRD GOLD |
| 2005 to 2009 | 1 | 77.83 (76.90 - 78.77) |  |  |
| 2010 to 2014 | 1 | 80.65 (79.79 - 81.53) |  |  |
| 2015 to 2019 | 1 | 80.10 (79.00 - 81.23) |  |  |
| 2020 to 2021 | 1 | 76.90 (74.57 - 79.29) |  |  |
| 2000 to 2004 | 5 | 52.85 (51.34 - 54.40) | Both | CPRD Aurum |
| 2005 to 2009 | 5 | 49.72 (48.61 - 50.86) |  |  |
| 2010 to 2014 | 5 | 50.84 (48.62 - 53.16) |  |  |
| 2015 to 2019 | 5 | 54.73 (53.64 - 55.84) |  |  |
| 2000 to 2004 | 5 | 50.01 (47.33 - 52.84) |  | CPRD GOLD |
| 2005 to 2009 | 5 | 50.35 (48.83 - 51.92) |  |  |
| 2010 to 2014 | 5 | 53.69 (52.22 - 55.20) |  |  |
| 2015 to 2019 | 5 | 53.40 (51.64 - 55.22) |  |  |
| 2000 to 2004 | 5 | 57.07 (55.36 - 58.83) | Female | CPRD Aurum |
| 2005 to 2009 | 5 | 50.77 (49.11 - 52.49) |  |  |
| 2010 to 2014 | 5 | 49.44 (43.88 - 55.71) |  |  |
| 2015 to 2019 | 5 | 55.74 (54.14 - 57.39) |  |  |
| 2000 to 2004 | 5 | 51.78 (47.88 - 56.00) |  | CPRD GOLD |
| 2005 to 2009 | 5 | 50.61 (48.06 - 53.30) |  |  |
| 2010 to 2014 | 5 | 53.64 (51.44 - 55.93) |  |  |
| 2015 to 2019 | 5 | 55.79 (53.33 - 58.36) |  |  |
| 2000 to 2004 | 5 | 49.49 (47.22 - 51.88) | Male | CPRD Aurum |
| 2005 to 2009 | 5 | 48.86 (47.38 - 50.38) |  |  |
| 2010 to 2014 | 5 | 51.38 (49.82 - 52.98) |  |  |
| 2015 to 2019 | 5 | 53.92 (52.44 - 55.43) |  |  |
| 2000 to 2004 | 5 | 48.73 (45.13 - 52.60) |  | CPRD GOLD |
| 2005 to 2009 | 5 | 50.15 (48.35 - 52.01) |  |  |
| 2010 to 2014 | 5 | 53.73 (51.78 - 55.76) |  |  |
| 2015 to 2019 | 5 | 51.52 (49.07 - 54.09) |  |  |

CI: confidence interval
